# PRX-3140, a 5-HT4 Partial Agonist and Sigma-1 Agonist/Antagonist, Modulates Glucocorticoid Insulin Suppression and Cortisol Levels

**DOI:** 10.1101/2024.12.04.24318244

**Authors:** B.L. Hood, J.P. McLaughlin, A.R. Alleyne, J.S. Thinschmidt, S.W. Harden, C.J. Frazier, J.D. Talton

**Author notes:** **Correspondance:** James D. Talton, Ph.D. Nanopharmaceutics, Inc. 14120 NW 126 Terrace, Alachua, FL 32615.

## Abstract

PRX-3140 is a partial agonist to the 5-hydroxytryptamine receptor 4 (5-HT4) and a ligand for the sigma-1 (S1R) and sigma-2 (S2R) receptors. Although few publications have inferred S1R agonists/antagonists modulate blood glucose, Di et.al (2017) reported S1R deficiency in knockout mice impacted regulation of the hypothalamic-pituitary-adrenocortical (HPA) axis, with a dexamethasone-induced reduction in level of corticosterone markedly attenuated in S1R −/− knockout mice, implicating S1R in feedback response to the HPA axis. The hypothesis that S1R deficiency causes down-regulation of the glucocorticoid receptor (GR) and attenuates GR-mediated feedback inhibition of HPA axis, as well as stress response of HPA axis, suggest that the inverse, the activation of S1R under normal conditions, may modulate glucocorticoid insulin suppression (as a direct S1R-GR effect) as well as cortisol levels (producing HPA axis feedback inhibition). In the present study, coadministration of 10 µM PRX-3140 with 100 nM cortisol significantly increased insulin release (to 74.8 ng/ml, P-value <0.0001). Similar effects were observed when cells were exposed to dexamethasone (Dex), with 10 µM PRX-3140 and 10 nM Dex producing 1.87-fold significantly more insulin than 10 nM Dex alone. Daily glucose concentrations in the 14-day clinical study (NCT00384423) of PRX-3140 demonstrate a reduction for 10 mg once-daily at days 1, 7, 10, and 15. Urine free cortisol levels at 10, 30, 100 and 200 mg dose levels of PRX-3140 demonstrated a larger reduction at 7 and 14 days compared to placebo. As an agonist of S1R that acts as a chaperone of GR, PRX-3140 has demonstrated GR modulating effects in INS-1 cells and in 14-day clinical studies in healthy adults with low incidence of side effects. The results of the present study suggest that S1R activation, with PRX-3140 and NP-18-2 S1R agonists, modulates glucocorticoid insulin suppression and cortisol levels.

## 1 Introduction

PRX-3140 is a partial agonist to the 5-hydroxytryptamine receptor 4 (5-HT4) and a ligand for the sigma-1 (S1R) and sigma-2 (S2R) receptors that has shown promise in preclinical and clinical studies for its potential therapeutic effects in cognitive disorders, particularly Alzheimer’s disease. [Johnson-2012]. By modulating S1R activity, PRX-3140 may enhance neuroprotective mechanisms, improve synaptic function, and promote neuronal health, thereby addressing neurodegenerative conditions associated with cognitive deficits [Nguyen-2017]. Studies have indicated that S1R agonists can positively influence neurotransmitter systems, including acetylcholine (ACh), which is crucial for memory and learning [van Waarde-2011]. In male Long-Evans rats, PRX-3140 intraperitoneal (IP) injection increased ACh output in the hippocampal formation under “resting” and behavioral testing conditions [Johnson-2012]. Furthermore, early clinical trials have suggested that PRX-3140 is well-tolerated and may lead to improvements in cognitive function in patients with mild to moderate Alzheimer’s disease. Continued research into PRX-3140 could provide insights into its efficacy and safety, as well as its broader implications for treating cognitive impairments.

Three Phase 1 and two Phase 2 trials in 248 subjects have been conducted with PRX-3140 dosed orally at 5 to 250 mg daily up to two years. Most adverse events were mild to moderate in severity, and occurred primarily in the higher dose groups, including dizziness, postural dizziness, headache, abnormal dreams, nausea, diarrhea, dyspepsia, decreased appetite, and somnolence. Whereas earlier 5-HT4 agonists (e.g., cisapride and tegaserod) have been associated with cardiovascular adverse events, including hERG (cardiac potassium) channel effects (QT prolongation), no cardiovascular safety concerns were reported for PRX-3140. In dose escalation studies, improvement in cognitive scores in subjects with mild Alzheimer’s Disease were observed from 5 to 50 mg delivered orally once a day. In a two-week clinical study (NCT00384423), clinical laboratory measurements demonstrated modulation of glucose levels and cortisol at low doses of PRX-3140. However, post-hoc analysis was not performed to address this effect.

Drugs with S1R activity, such as haloperidol [Walter-2006], dextromethorphan [Marquard-2014], and pentazocine [Kavitha-1999] have been shown to have effects on glucose levels. Only a few publications have inferred that S1R agonists/antagonists modulate blood glucose levels [Paniagua-2016]. In 2017, Di et.al reported S1R deficiency in knockout mice impacted regulation of the hypothalamic-pituitary-adrenocortical (HPA) axis such that acute mild restraint stress (AMRS) induced a higher and more sustainable increase in activity of HPA axis [Di-2017]. In another experiment, Dexamethasone (Dex)-induced reduction in corticosterone levels was markedly attenuated in S1R −/− knockout mice, implicating S1R in glucocorticoid mediated feedback inhibition of the HPA axis. The hypothesis that S1R deficiency causes down-regulation of the glucocorticoid receptor (GR) and attenuates GR-mediated feedback inhibition of HPA axis, as well as stress response of HPA axis, suggest that the inverse, activation of S1R under normal conditions, may modulate glucocorticoid mediated insulin suppression (direct S1R-GR effect) as well as cortisol levels (HPA axis feedback inhibition). The present study investigated the relationship between PRX-3140 and other S1R agonists to glucocorticoid insulin suppression and cortisol levels to obtain a better understanding of these direct and indirect mechanisms compared to 5-HT4 activity.

## 2 Materials and Methods

### 2.1 Chemicals

S1R ligands 6,7-Dihydro-4-hydroxy-7-isopropyl-6-oxo-N-(3-(piperidin-1-yl)propyl)thieno[2,3-b]pyridine-5-carboxamide potassium salt (PRX-3140), N-{3-[4-(4-cyclohexylmethanesulfonyl aminobutyl)-piperazin-1-yl]phenyl} acetamide hydrochloride (NP-2), and 5-((4-(6-chlorothieno[2,3-d]pyrimidin-4-ylamino)piperidine-1-yl)methyl)-2-fluorobenzonitrile monofumarate (NP-3) were all synthesized in-house (Alchem Laboratories Corp.). All other reagents were purchased from Sigma (St. Louis, MO) unless otherwise noted.

### 2.2 Receptor Binding Assays

*In vitro* radioligand competition receptor binding assays were performed to evaluate the pharmacologic profiles of PRX-3140, NP-2 and NP-3, including the S1R, S2R, a panel of serotonin receptor subtype assays (5-HT1A, 5-HT2B, and 5-HT4) and other off-target receptor subtypes. Standard receptor binding methods were employed and the majority of assays were performed using a human recombinant receptor. Where a human assay system was unavailable, receptor binding was performed in tissue from mouse, rat, guinea pig, chicken, or bovine source.

Human T lymphocyte Jurkat cells and guinea pig cerebral cortex were used to prepare S1R in HEPES/Tris buffer pH 7.4. PRX-3140 was incubated with 2 and 15 nM [^3^H] Pentazocine for 120-150 minutes. Non-specific binding is estimated in the presence of 10 µM Haloperidol. Membranes were filtered and washed, and the filters then counted in a beta scintillation counter to quantify total binding of [^3^H] Pentazocine, with specific binding then calculated by subtracting non-specific binding. Inhibition constants (Ki) values were calculated using the equation of Cheng and Prusoff [Cheng-1973] using the observed IC50 of the tested compound, the concentration of radioligand, and the historical values for the KD of the ligand (obtained experimentally at Eurofins Panlabs, Inc.) ± the standard error of the mean (SEM).

### 2.3 Acute Brain Slice Preparation and Electrophysiology Studies

All procedures performed on live animals as described below were reviewed and approved by the Institutional Animal Care and Use Committee at the University of Florida. Animals were deeply anesthetized using a ketamine/xylazine cocktail (0.1 ml of 10% ketamine and 0.05 ml of 2% xylazine). Brains were quickly extracted and submerged into ice-cold sucrose-laden normal artificial cerebrospinal fluid (ACSF; 206 mM sucrose, 10 mM d-glucose, 1 mM MgSO_4_, 2 mM KCl, 1.25 mM NaH_2_PO_4_, 1 mM CaCl_2_, and 25 mM NaHCO_3_). A VT1000s vibratome (Leica Microsystems, Buffalo Grove, IL) was used to make 300 μm coronal sections that included the nucleus accumbens (NAc). Sections were maintained at 37 °C in normal ACSF (126 mM NaCl, 11 mM d-glucose, 1.5 mM MgSO_4_, 3 mM KCl, 1.2 mM NaH_2_PO_4_, 2.4 mM CaCl_2_, and 25mM NaHCO_3_) and subsequently equilibrated to room temperature for a minimum of 30 minutes prior to experimental use. Importantly, normal ACSF was used for incubation rather than a low-Ca^2+^ ACSF (1 mM CaCl_2_) as NAc MSNs (medium spiny neurons) do not regain normal AP firing function following prolonged submersion in the latter [Scala et al., 2018; Tapia et al., 2020]. All external solutions were saturated with 95% O_2_ / 5% CO_2_ and had a pH of 7.3.

Brain slices used in whole-cell patch clamp electrophysiology studies were transferred to a recording chamber continuously perfused at 2 mL/min with oxygenated ACSF maintained at 28°C. Picrotoxin (PTX, 100 μM), 6,7-dinitroquinoxaline-2,3-dione (DNWX, 20 μM), and (2R)-amino-5-phosphonovaleric acid (AP5, 40 μM) were added to ACSF to block fast synaptic currents mediated by GABA_A_ receptors, AMPA/kainate receptors, and NMDA receptors, respectively. An Olympus BX51WI stereomicroscope supporting infrared differential interference contrast microscopy (IR-DIC) was used to visualize cells.

Patch pipettes (borosilicate glass capillaries 1.5 mm/0.8 mm, Sutter Instrument Company) were made using a Flaming/Brown-type pipette puller (Sutter Instrument, P-07), and had an with an open tip resistance of 4-6 MΩ when filled with a K-gluconate based internal solution that contained in mM: 2 MgCl_2_, 0.5 EGTA, 10 HEPES, 115 K-gluconate, 10 phosphocreatine, 4 Na_2_-ATP, 0.4 Na_3_-GTP, and 5 KCl adjusted. This solution was adjusted to pH 7.25 using KOH and volume adjusted to 295 mOsm. Whole-cell recordings were made using a Multiclamp 700B amplifier, Digidata 1440A digitizer, and Clampex 10.7 software (Molecular Devices). All whole-cell data were sampled at 20 kHz and low-pass filtered at 2 kHz. Cells were identified as NAc MSNs based on location (proximity to the corpus collosum) and resting membrane potential (≤ -70 mV) [Belleau and Warren, 2000; Cao et al., 2016; Willett et al., 2018; Aceto et al., 2022].

Analysis of electrophysiological recordings was performed with custom software written using Python 3.10 and the pyABF package [Harden, 2022] and in OriginC (OriginLab Corporation, Northampton, MA). Data presented were not corrected for the liquid junction potential. Cells were excluded from analysis that had a resting membrane potential > -70 mV, that did not survive through experiment completion, or that experienced a sudden change in seal quality during recording.

Frequency of evoked action potentials (APs) was recorded from NAc MSNs in current-clamp configuration. Evoked APs were measured during a 4 second current injection starting from -100 pA with a 20 pA increase per sweep, every 10 seconds. The resting membrane potential (RMP) was measured at the beginning of each sweep as the average voltage observed over a 3-second period with I = 0, when cells were quiescent.

### 2.4 INS-1 832/13 Rat Insulinoma Cell Line

The INS-1 832/13 rat insulinoma cell line was purchased from Millipore (Temecula, CA) and cultured according to manufacturer instructions. Briefly, cells were thawed and expanded in RPMI-1640 supplemented with L-Glutamine and 25mM HEPES (Corning, Manassas, VA), 1 mM sodium pyruvate, 10 mM HEPES, 0.05 mM β-mercaptoethanol, 100 U/ml penicillin (Gibco, Grand Island, NY), 100 μg/ml streptomycin (Gibco, Grand Island, NY), and 10% EmbryoMax^®^ ES Cell Qualified FBS at 37°C, 5% CO_2_, 95% humidity. After 2 days in culture, medium is exchanged with fresh medium containing G-418 (Gibco, Grand Island, New York) at 0.3 mg/ml for selective pressure. When the cells were confluent, cells were subcultured with a 1 to 6 split. For subculturing, cells are detached using 0.25% Trypsin-EDTA (Gibco, Grand Island, NY). Passages 13 through 27 were used for the insulin release assays.

### 2.5 Insulin Secretion Assays

The insulin secretion assay was based on the Millipore manufacturer insert for the INS-1 832/13 cells as well as the original published paper [Hohmeier, et al, 2000]. Briefly, the INS-1 832/13 cells were seeded in Costar 24-well TC plates (Corning, NY) at a cell density of 5X10^5^ cells per well using the expansion media without G418. Cultures were then incubated for 48 hours 37°C, 5% CO_2_, 95% humidity to confluency. The media was then replaced with fresh media containing test compounds. The control wells were 2.5 mM glucose and 16.7 mM glucose with 0.1% DMSO. Plates were incubated for 18 hours (37°C, 5% CO_2_, 95% humidity), then wells were washed with HBSS twice (114 mmol/L NaCl, 4.7 mmol/L KCl, 1.2 mmol/L KH_2_PO_4_,1.16 mmol/L MgSO_4_, 20 mmol/L HEPES, 2.5 mmol/L CaCl_2_, 25.5 mmol/L NaHCO_3_, and 0.2% bovine serum albumin, pH 7.2). Next HBSS containing 2.5 mM glucose was added and incubated 1 hour (37°C, 5% CO_2_, 95% humidity). Cells were then washed twice with HBSS and followed by addition of HBSS with 2.5 mM or 16.7 mM glucose, as well as test compounds diluted in 16.7 mM glucose. Note that DMSO was supplemented in the control wells to maintain a consistent DMSO concentration (0.1%) in all wells. After a final incubation for 2 hours (37°C, 5% CO_2_, 95% humidity), the supernatant was removed to measure the concentration of insulin using the ALPCO Insulin Rodent (Mouse/Rat) Chemiluminescence ELISA (ALPCO, Salem NH).

### 2.6 Insulin ELISA Assay

Insulin secretion was evaluated by using the ALPCO Insulin Rodent (Mouse/Rat) Chemiluminescence ELISA (ALPCO, Salem NH). Briefly, 75 µl of working strength conjugate was dispensed to each well pre-coated with a monoclonal antibody specific to insulin. Next, 5 μl of the standards or test samples were dispensed in duplicate. The plate was sealed and incubated at room temperature for 2 hours at 700 rpm on a microplate shaker. After the incubation, the content of the wells was decanted and washed 6 times with 350 µl of working strength wash buffer. After each wash, the plate was firmly tapped on absorbent paper to remove residual liquid. After the last wash, 100 µl of working chemiluminescent substrate was dispensed into each well and incubated 5 minutes at room temperature. Then, 80 µl were transferred to a pureGrade™ S white plate (Brandtech Scientific, Dawsonville, GA). The plate is immediately read on the PerkinElmer Envision multi-mode plate reader in ultra-sensitive luminescent mode.

### 2.7 ATPLite Cytotoxicity Assay

Test compounds were diluted in DMSO. Ten nanoliters of each compound or combination of compounds were dispensed to the 6 designated test wells of a Corning 1536 well TC white plate (Corning, Kennebunk, ME) using a Labcyte/Beckman Coulter Echo 555 acoustic dispenser. For a positive control for 100% cell death, 10 nl of 10 mM MG-132 was dispensed to the positive control wells of the assay plate (final assay concentration 25 µM) and for the negative controls (no cell death), 10 nl of DMSO was dispensed to negative control wells of the assay plate. Next, the cells were detached from the flask, centrifuged, counted on a Cytometer T4 (Nexcelom Biosciences, Lawrence, MA) and resuspended in growth medium at a density of 400 cells per 4 µl. Then the cells were dispensed to the assay plate using the Thermo-Scientific™ Multidrop™ Combi Reagent Dispenser (4 µl of cells in each well). The plate was centrifuged at 800rpm for 15 seconds and the clear lid was replaced with a Kalypsys lid (Kalypsys, Inc. San Diego, CA) to reduce edge effects and allow the even distribution of CO_2_ to all wells of the plate. The plate was incubated for 48 hours at 37°C, 5% CO_2_, 95% humidity. After the incubation, 4 µl of ATPlite 1step reagent (Revvity, Waltham, MA) was dispensed into the wells using a Beckman Coulter BioRAPTR FRD. The assay plate was centrifuged at 1000rpm for 1 minute and incubated in the dark for 15 minutes at room temperature. The signal was then read on a Perkin Elmer Envision multi-mode plate reader in ultra-sensitive luminescence mode.

### 2.8 Clinical Studies

Data from a 14-day clinical study (NCT00384423) with PRX-3140 were analyzed (Quintiles. Inc. CRO). Each study protocol was approved by their respective investigational review board or human subjects committee. Healthy volunteers were initially enrolled with six subjects completing all treatments. Serum concentrations of PRX-3140 were determined using a validated liquid chromatography coupled with tandem mass spectrometry (LC/MS/MS) method. Serum concentration-time data were analyzed by noncompartmental methods using WinNonlin^®^ Professional. The first cohort to evaluate the safety, tolerability and pharmacokinetics of PRX-3140 was conducted in 32 healthy adult male and female volunteers from 18 to 45 years of age. Each subject was administered an oral dose (capsules) of PRX-3140 or placebo once daily for 14 days. Eight (8) subjects were randomly assigned to each of 4 sequential dosing cohorts and received either 10 mg, 30 mg, 100 mg, or 200 mg of PRX-3140 or matched placebo. If a dose regimen was found to be safe and well tolerated, then the succeeding group of 8 different subjects received the next higher dose of PRX-3140 (N = 6) or placebo (N = 2) for 14 days. Dose escalation was dependent on safety parameters (physical examination findings, vital signs, adverse events (AEs), electrocardiograms (ECGs), and clinical laboratory values including glucose and cortisol). Urine free cortisol was measured by LC-MS/MS from the first morning void urine samples collected at baseline (predose on Day 1) and predose on Days 7 and 14. Continuous glucose monitoring (CGM) was applied in a single patient with placebo compared to 10 mg PRX-3140. Clinical pharmacokinetic (PK) and pharmacodynamic (PD) sampling was performed. Subjects were monitored closely throughout the study.

### 2.9 Statistical Analysis

Within groups, a paired Student’s t-test was used to evaluate the effect relative to the baseline mean. Across groups, an unpaired Student’s t-test was used to compare data at matching time points. Differences were considered significant where *p* ≤ 0.05.

For patch clamp studies, data are reported as mean ± SEM. Comparisons between groups were made with a one-way ANOVA in OriginPro (Originlab, Northampton, MA) using Sidak-Holm post-hoc tests when indicated.

INS-1 832/13 rat insulinoma cell line insulin release data analysis was done using GraphPad Prism Box and Whiskers plots graphed using the Tukey Boxplots analysis. The software calculates the IQR which is the inter-quartile distance (the difference between the 25th and 75th percentile). Then adds the 75th percentile to 1.5 times the IQR. If this value is greater or equal to the largest value in the data set, an upper whisker is drawn at the largest value. If this value is less than the largest value, the upper whisker is set at the largest value less than the 75th percentile plus 1.5 times IQR. Any values that are greater than this value are plotted as individual points and are considered outliers for our study. For the lower whisker, the software calculates the 25th percentile minus 1.5 times IQR. If this value is less than the smallest data value, the whisker is drawn at to the smallest data point. If the value is greater than the smallest data point, the whisker is set at the lowest value greater than the 25th percentile minus 1.5 times IQR. Any values that are less than this value are drawn as individual points (again, these are considered outliers for our study). The GraphPad Prism software also was used to determine P values for the Student’s T-tests. The percent cell death in the cytotoxicity assays was determined using GeneData software (GeneData, Basel, Switzerland) which normalizes the data in reference to the control values.

Pharmacokinetic analysis was performed on patients who received active study treatment. AUC24 and Cmax values reported as geometric mean (CV%). T1/2, glucose concentrations and urine free cortisol are expressed as arithmetic mean (SD).

## 3 Results

### 3.1 5-HT and S1R Receptor Binding

PRX-3140 was evaluated in a panel of serotonin receptor subtype assays to establish the selectivity for 5-HT4 binding sites. PRX-3140 shows high affinity (Ki = 22nM) and selectivity for 5-HT4R with more than 100-fold difference in affinities compared with all other 5-HT receptors tested. PRX-3140 also has good affinity for the S1R (Ki = 79-160nM) and S2R (Ki = 40-100nM) in receptor binding assays. The binding data for the 5-HT1A, 5-HT2B, 5-HT4, S1R, S2R activity is listed in Table 1.

**Table 1:** Receptor Binding affinity of PRX-3140, NP-18-2 and NP-18-3. Binding Ki values for 5-HT1A, 5HT2B, 5-HT4, S1R and S2R receptors were determined by using human recombinant receptor or tissue from mouse, rat, guinea pig, chicken, or bovine source. For PRX-3140, Ki values are reported as the geometric mean and S.E.M. based on four replicates (S.E.M. values expressed are shown in parentheses.)

| Ki | 5-HT1A | 5-HT2B | 5-HT4 | S1R | S2R |
| --- | --- | --- | --- | --- | --- |
| PRX-3140 | >5 uM | >5 uM | 14.3 nM (5.4) | 10.0 nM (3.3) | 36.1 nM (16.1) |
| NP-18-2 | 5-11 nM | >5 uM | >5 uM | 100 nM | >2 uM |
| NP-18-3 | 100 nM | 3.4 nM | >5 uM | 110 nM | 42 nM |

NP-18-2 is a selective full agonist to the 5-HT1A and a ligand for the S1R. NP-18-2 is a high affinity (Ki = 5.1-17 nM) 5-HT1A agonist in radioligand binding assays. NP-18-2 also demonstrates affinity for the S1R (Ki = 100 nM) in radioligand binding assays (Table 1).

NP-18-3 is a selective full antagonist to the 5-HT2B receptor and a ligand for the S1R. NP-18-3 is a highly selective for 5-HT2B (Ki=3.4 nM) with more than 500-fold differences in affinity for 5-HT2BR compared with all other 5-HT receptor subtypes tested except for the 5-HT1A receptor (5-HT1A, Ki = 100nM). NP-18-2 also demonstrates binding to the S2R (Ki = 42 nM) and S1R (Ki = 100 nM) in radioligand binding assays. NP-18-3 affinities for receptors are listed in Table 1.

### 3.2 PRX-3140 Reduces Excitability of Medium Spiny Neurons in the Nucleus Accumbens

In order to determine whether PRX-3140 directly interacts with S1R in vivo, we evaluated its ability to impact the intrinsic excitability of medium spiny neurons (MSNs) in the NAc. Specifically, we performed whole-cell patch clamp recordings from visually identified MSNs in acute tissue slices through the NAc (see methods). MSNs were injected with a series of current pulses ranging from -120 to 800 pA, and action potential frequency in response to each stimulus was measured in current clamp. Our results revealed a concentration-dependent decrease in maximum action potential frequency observed in response to current injection (Figure 1, one-way ANOVA, F_4, 24_ = 17.6, p < 0.0001). Additional post-hoc tests revealed a significant effect of PRX-2140 vs. control at 10 µM and 100 µM (p<0.0001 in both cases), but not at 500 nM or 1 µM. Overall, these effects of PRX-3140 are consistent with the reported results of other known S1R interacting compounds as described in greater detail in [Alleyne-2023].

**Figure 1:**
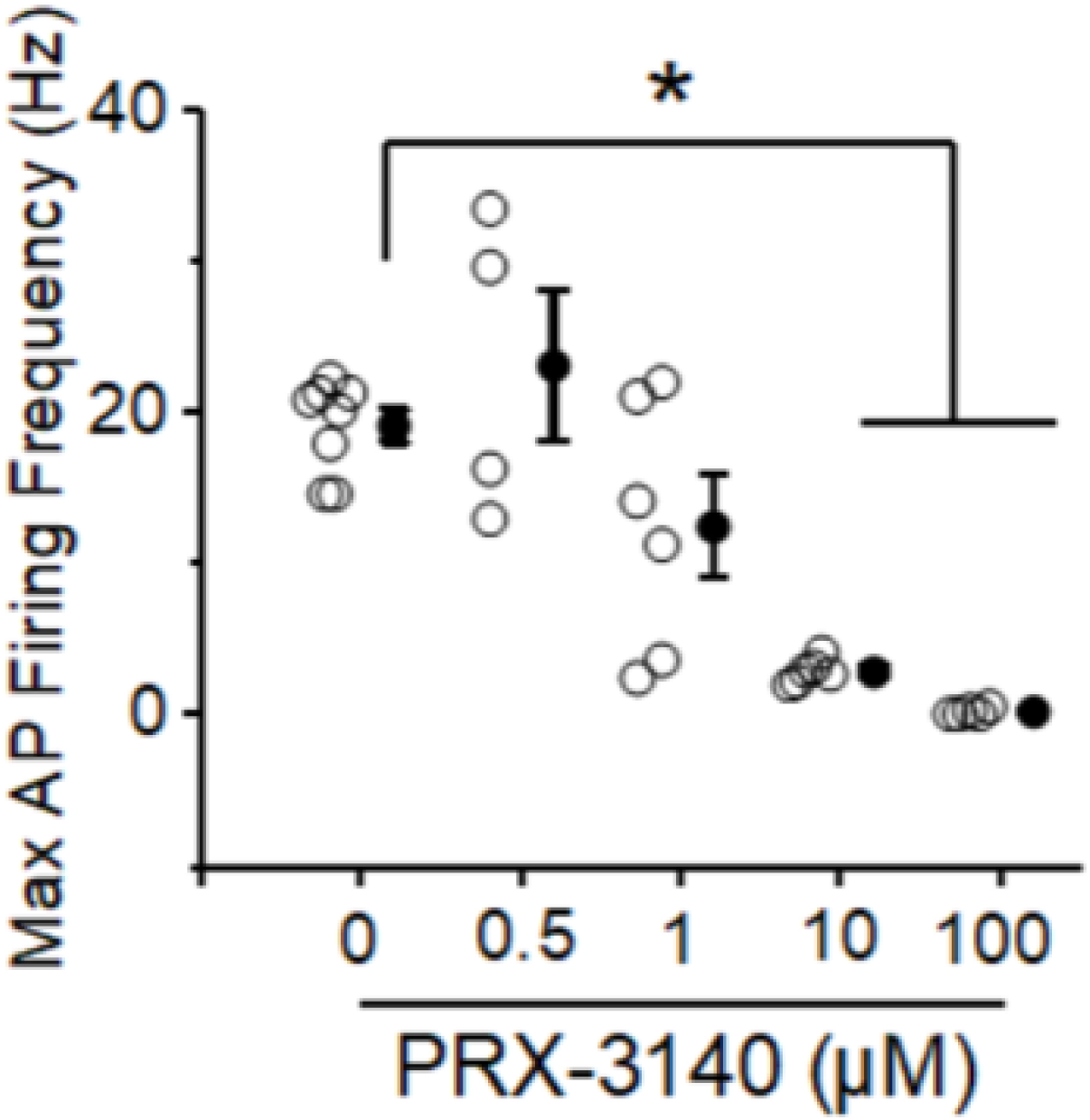
PRX-3140 reduces maximum firing rate of NAc MSNs in a concentration-dependent manner. *Ex vivo* application of PRX-3140 produces a dose-dependent reduction in maximum firing rate observed in NAc MSNs in response to a transient (4 second) depolarizing current injection. Open circles are individual data points and closed circles are the mean at each concentration concentration (error bars +/- SD.) A one-way-ANOVA on these data reveals a statistically significant main effect of PRX-3140 (see results for additional details). Asterisks indicate statistical significance on pairwise comparisons from post-hoc tests.

### 3.3 Insulin Release Modulation by 5-HT/S1R Agonists in INS-1 832/13 Rat Insulinoma Cells

To examine the effect of PRX-3140 on insulin production in INS-1 832/13 cells, the cells were exposed for 18 hours to varying concentrations of PRX-3140 and then starved for 1 hour with low glucose (2.5 mM glucose). The test compound/compounds were then added back to the cells diluted in 16.7 mM glucose. The addition of 16.7 mM glucose with 10 µM PRX-3140 or 10 nM dehydroepiandrosterone (DHEA) showed a slight increase of insulin release in these cells (138.2 and 138.0 ng/ml, respectively compared to 113.8 ng/ml for 16.7 mM glucose with no compound) but the difference was not significant (Figure 2). In our studies, 10 µM dehydroepiandrosterone sulfate (DHEA-S) slightly decreased the insulin release (107.8 ng/ml compared to 112 ng/ml glucose alone).

**Figure 2:**
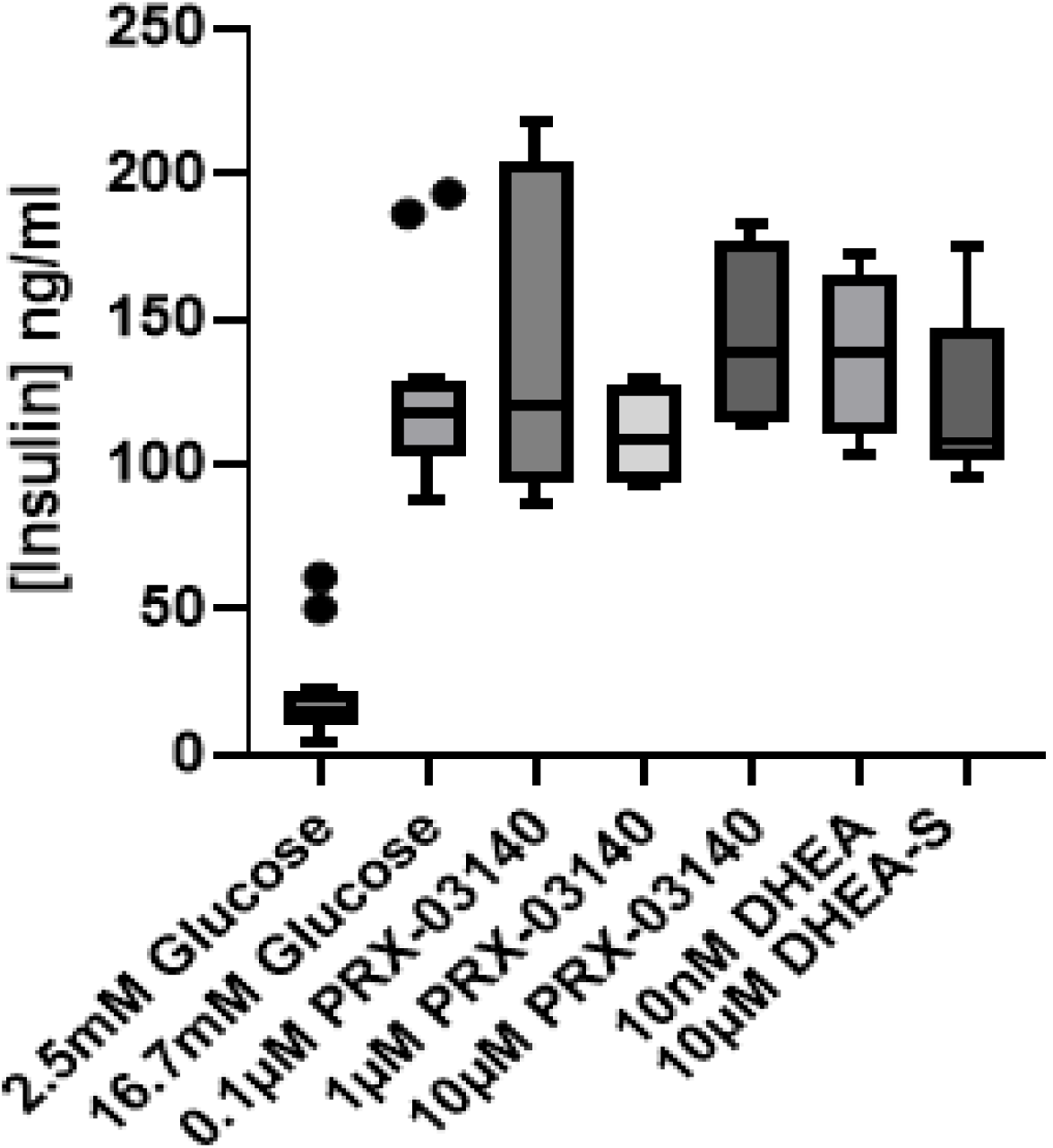
Response of INS-1 832/13 Cells to PRX-1340, DHEA and DHEA-S. After an 18 hours incubation with the test compounds, cells were washed, starved for 1 hour with 2.5 mM glucose and then exposed for 2 hours with the test compounds diluted in 16.7 mM glucose. The insulin content in the supernatant was determined by ELISA. Each condition is representative of an N of 4 (solid circles denote outliers.) Data was analyzed using GraphPad Prism using the Box and Whiskers plot.

To examine the effect of endogenous glucocorticoids on insulin production in INS-1 832/13 cells, the cells were exposed to cortisol and corticosterone at physiologically relevant concentrations for 18 hours, washed with HBSS twice, starved for 1 hour with low glucose (2.5 mM glucose) and then exposed to the glucocorticoids diluted in 16.7 mM glucose for 2 hours triggering the insulin release. The amount of insulin release was then determined using the insulin ELISA assay (Figure 3). In these studies, 100 nM cortisol diluted in 16.7 mM glucose significantly reduced insulin release (median value 48.8 ng/ml) compared to 16.7 mM glucose alone (median value 117.3 ng/ml; p<0.0001; Student’s T-test). As expected, 500 nM cortisol had more of an effect on insulin release with a median value of 37 ng/ml. Corticosterone (100 nM in 16.7 mM glucose) also significantly decreased the insulin release giving a median value 78.2 ng/ml (P value = 0.0011). If 10 µM PRX-3140 was co-administered with the 100 nM cortisol, the insulin release was significantly increased (median value of 74.8 ng/ml, P<0.0001). If 1 µM DHEA-S was co-administered with 100 nM cortisol, there was no difference between this combination compared to 100 nM cortisol alone. If a combination of 100 nM cortisol plus 10 nM DHEA was added to the cells, the insulin release dropped to 37.0 ng/ml which is significantly different from 100 nM cortisol alone (P value = 0.0015). The combination of 500 nM Cortisol plus 10 nM DHEA had a median value of 33.2 ng/ml which is statistically the same as 500 nM cortisol (30.4 ng/ml).

**Figure 3:**
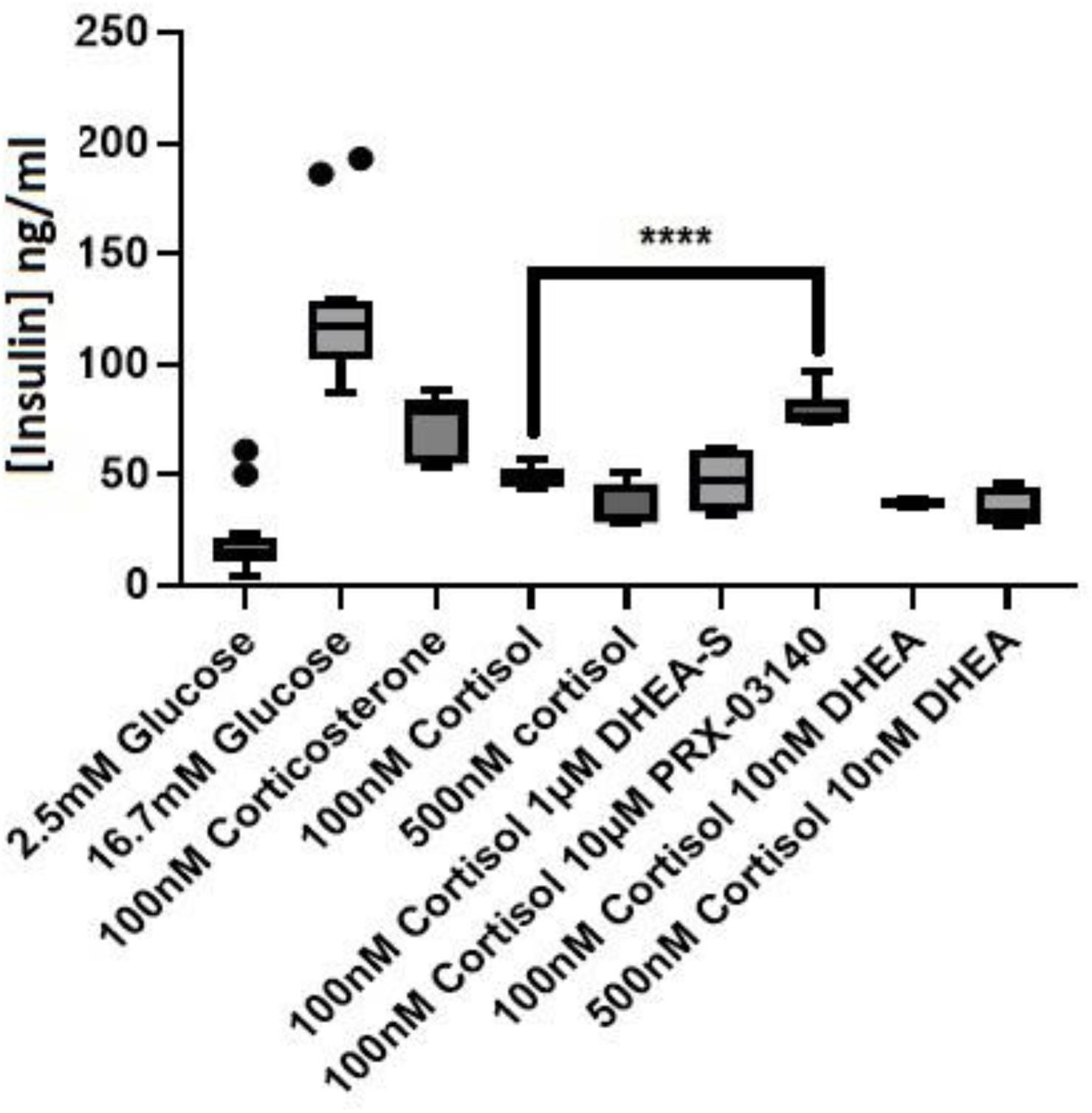
Response of INS-1 832/13 Cells to Corticosterone, Cortisol, Cortisol with PRX-3140, DHEA and DHEA-S. After an 18 hours incubation with the test compounds, cells were washed, starved for 1 hour with 2.5 mM glucose and then exposed for 2 hours with the test compounds diluted in 16.7 mM glucose. The insulin content in the supernatant was determined by ELISA. Each condition was tested at least 4 times. Data was analyzed using GraphPad Prism using the Box and Whiskers plot (solid circles denote outliers.) **** Comparison of 100 nM cortisol to 100nM cortisol with 10 µM PRX-3140; P<0.0001; Student’s T test.

To examine the effect of a highly potent glucocorticoid, dexamethasone (Dex) was tested in INS-1 832/13 cells. As described previously, chronic exposure to the compound/compounds for 18 hours was followed by starvation with 2.5 mM glucose and stimulation with 16.7 mM glucose with and without compounds. Then, insulin released was measured in the insulin ELISA. The results are shown in Figure 4. The amount of insulin produced with 10 nM Dex in the wells was 52.5 ng/ml while 100 nM Dex yielded 38.6 ng/ml of insulin. If 10 µM PRX-3140 was included with the 10 nM Dex, significantly more insulin was produced (98.3 ng/ml, P value = 0.0124). When 10 µM PRX-3140 was combined with 100 nM Dex, the amount of insulin produced was 62.6 ng/ml which was greater than the amount with 100 nM Dex alone but it was not considered significant when the T-test analysis was performed. NP-18-2 released 138.15 ng/ml which was almost identical to PRX-3140 (138.20 ng/ml). However, when combined with 10 nM Dex, NP-18-2 did not recover the insulin release to the extent that PRX-3140 did (76.4 ng/ml vs 98.3 ng/ml, respectively). When NP-18-3 was dispensed to the cells, the median insulin production was 126.7 ng/ml. In combination with 10 nM Dex, NP-18-3 yielded only 40.7 ng/ml which was less than 10 nM Dex alone (52.4 ng/ml). However, the difference here was not significant. Cytotoxicity at 48 hours in INS-1 832/13 rat insulinoma cells was dose dependent based on steroid concentration similar to published studies [Suksri-2022]. All S1R agonists alone were not cytotoxic (data not shown).

**Figure 4:**
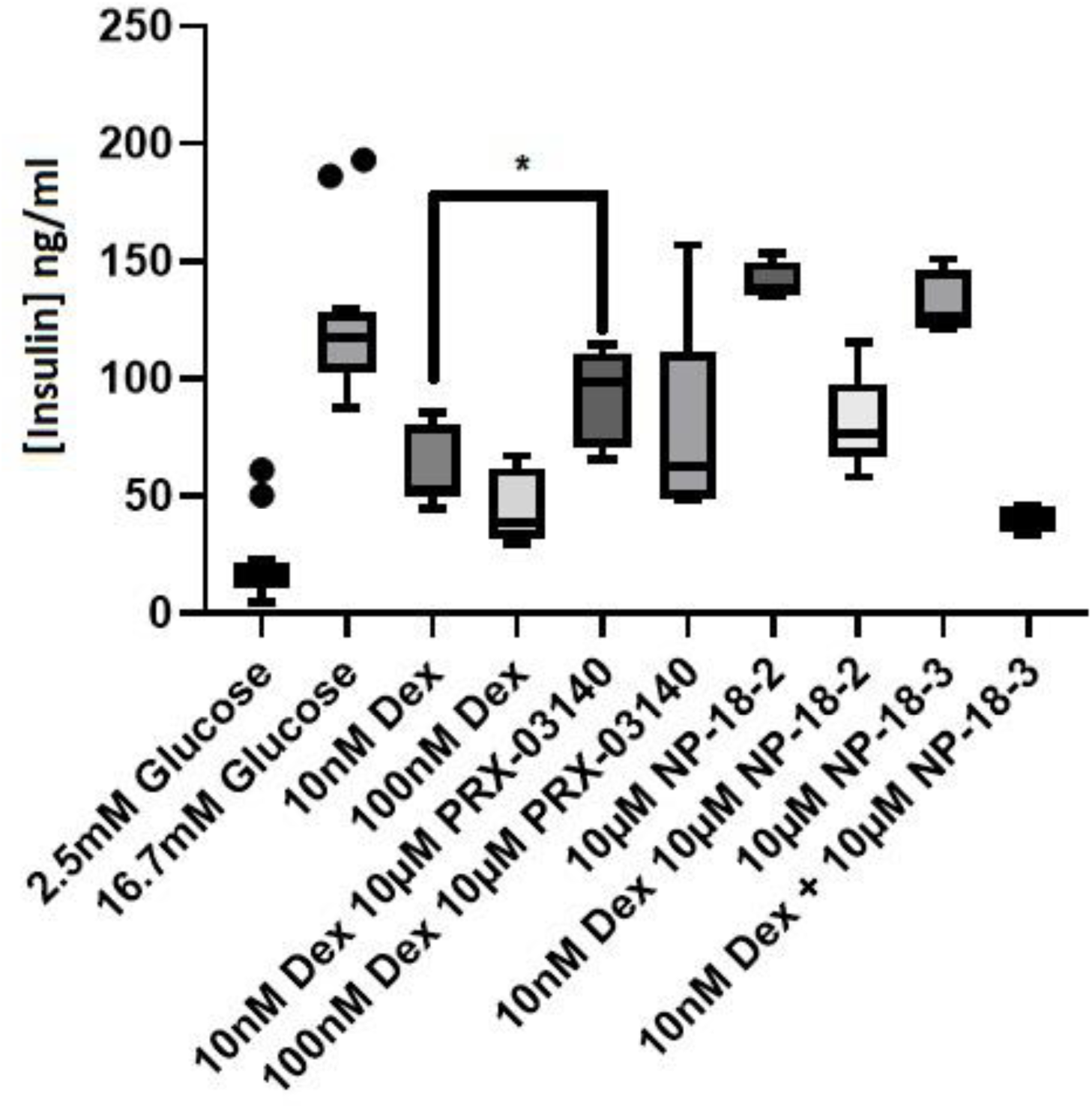
Response of INS-1 832/13 Cells to Dexamethasone and in Combination with NP-18-2, NP-18-3 or PRX-3140. After an 18 hours incubation with the test compounds, cells were washed, starved for 1 hour with 2.5 mM glucose and then exposed for 2 hours with the test compounds diluted in 16.7 mM glucose. The insulin content in the supernatant was determined by ELISA. Each condition was tested at least 4 times. Data was analyzed using GraphPad Prism using the Box and Whiskers plot (solid circles denote outliers.) * Comparison of 10nM Dex to 10 nM Dex with 10 µM PRX-3140; P=0.01; Student’s T test.

### 3.4 PRX-3140 Modulates Glucocorticoid Insulin Suppression and Cortisol Levels

Serum concentrations of PRX-3140 were detectable after administration of a single 5 mg dose and all other doses tested up to 16 hours and, in general, the mean serum concentrations of PRX-3140 increased with ascending single doses of 5 to 250 mg. The PK data suggested that PRX-3140 was absorbed at a relatively rapid rate (Tmax 1 to 2 hours) and the exposure showed moderate to high intersubject variability median values across this dose range studied but were generally dose-proportional. The Cmax values at the doses of 30 to 200 mg were less variable than those at the lower doses studied (5 and 10 mg). The mean half-life (T1/2) at doses of 50 to 250 mg was 8 to 14 hours, respectively, suggesting the feasibility of once daily dosing. The Cmax values at the doses of 25 to 250 mg (CVs 34-43%) were less variable than those at the lower doses studied. Following multiple oral dose administration of 10, 30, 100, or 200 mg PRX-3140 for 14 days, mean exposures (Cmax and AUC24) of PRX-3140 appeared to increase in a dose proportional manner. Dose-related increases in the mean trough serum concentration increases were evident throughout the treatment period and appeared to reach steady state by approximately Day 7 for the 10, 30, and 100 mg cohorts and Day 14 for the 200 mg PRX-3140 cohort. Non-compartmental PK parameters are presented in Table 2.

**Table 2.** Noncompartmental PK parameters for PRX-3140. Serum pharmacokinetic parameters include Tmax - Time to maximum serum concentration (h) obtained directly from the observed concentration versus time data on Days 1 and 14; Cmax = Maximum serum concentration (ng/mL) during the dosing interval obtained directly from the observed concentration versus time data on Days 1 and 14; AUC24 = Area under the serum concentration-time curve from time zero until 24 hours (ng·h/mL), calculated by linear trapezoidal summation; Cmin = Minimum serum concentration (ng/mL) during the dosing interval obtained directly from the observed concentration versus time data on Day 14; Cavg = Average steady-state serum concentration calculated as AUC24 divided by the dosing interval (24 h); T1/2 = Terminal half-life (h);. CV = coefficient of variation; SD = standard deviation. AUC24 and Cmax values reported as geometric mean (CV%) and T1/2 expressed as arithmetic mean (SD).

| PRX-3140 Dose | 5 mg | 10 mg | 30 mg | 50 mg | 100 mg | 200 mg |
| --- | --- | --- | --- | --- | --- | --- |
| <u>Day 1</u> |  |  |  |  |  |  |
| Tmax (h) | 2.0 | 1.5 | 1.5 | 1.0 | 1.5 | 1.0 |
| Cmax (ng/mL) | 12.1 (62) | 55 (36) | 76 (56) | 301.6 (43) | 641 (56) | 996 (38) |
| AUC24 (ng/mL*h) | 64 (89) | 159 (112) | 514 (82) | 1718 (156) | 3766 (73) | 4243 (46) |
| T1/2 (h) | 3.7 (2.4) | 4.2 (1.9) |  | 10.3 (9.8) |  |  |
| <u>Day 14 (Steady-state)</u> |  |  |  |  |  |  |
| Tmax (h) |  | 1.5 | 1.5 |  | 1.5 | 1.5 |
| AUC24 (ng/mL*h) |  | 533 (80) | 879 (102) |  | 5336 (47) | 11850 (39) |
| Cmax (ng/mL) |  | 77 (45) | 149 (47) |  | 729 (24) | 1573 (20) |
| Cmin (ng/mL) |  | 13.7 | 24.9 |  | 106.2 | 165.8 |
| Cavg (ng/mL) |  | 30.5 | 59.3 |  | 245.4 | 537.0 |
| T1/2 (h) |  | 12.7 (7.4) | 12.7 (14.4) |  | 10.2 (2.4) | 8.6 (3.0) |

Daily glucose concentrations in the 14-day study at 10, 30, 100 and 200 mg dose levels of PRX-3140 demonstrate a reduction for 10 mg once-daily at days 1, 7, 10, and 15 (Figure 5). Because of the small patient number (n=6 for PRX-3140 groups) and interpatient variability, this reduction was not statistically significant (Table 3.) Average daily glucose concentrations for placebo vs. 10mg PRX-3140 using CGM were 125 and 108 mg/dL, respectively, demonstrating a 15.6% reduction (data not shown). Out-of-range % > 140 mg/dL showed a reduction for 10mg PRX-3140 compared to placebo (4.1% vs. 17%, respectively), demonstrating an improvement in glycemic control.

**Figure 5:**
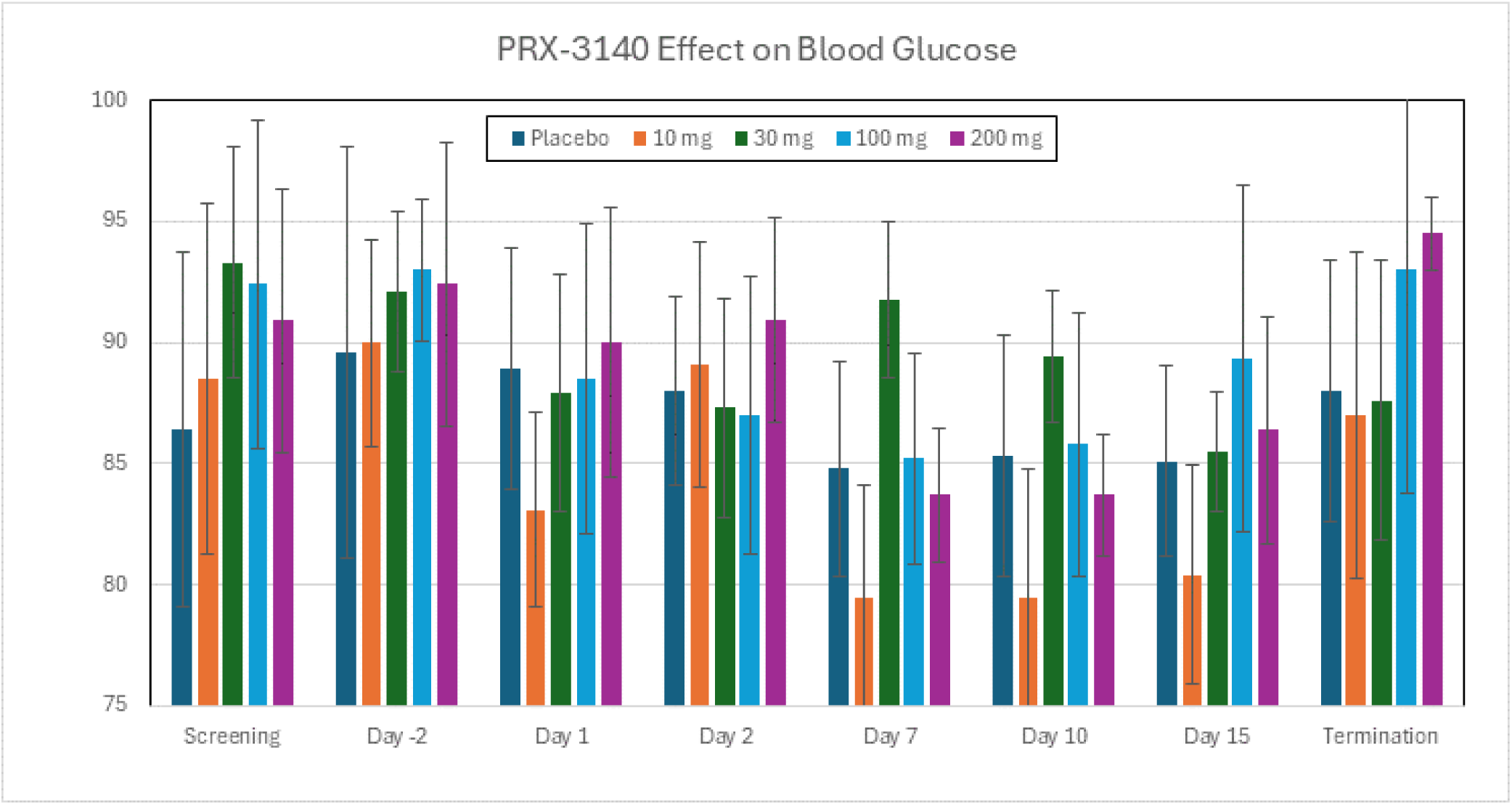
PRX-3140 effect on blood glucose. All laboratory assessments were performed at the clinical site’s certified laboratory utilizing that laboratory’s normal ranges. Blood samples were collected at Screening, Day -2, 1, 2, 7, 10, 15 and following termination.

**Table 3.**
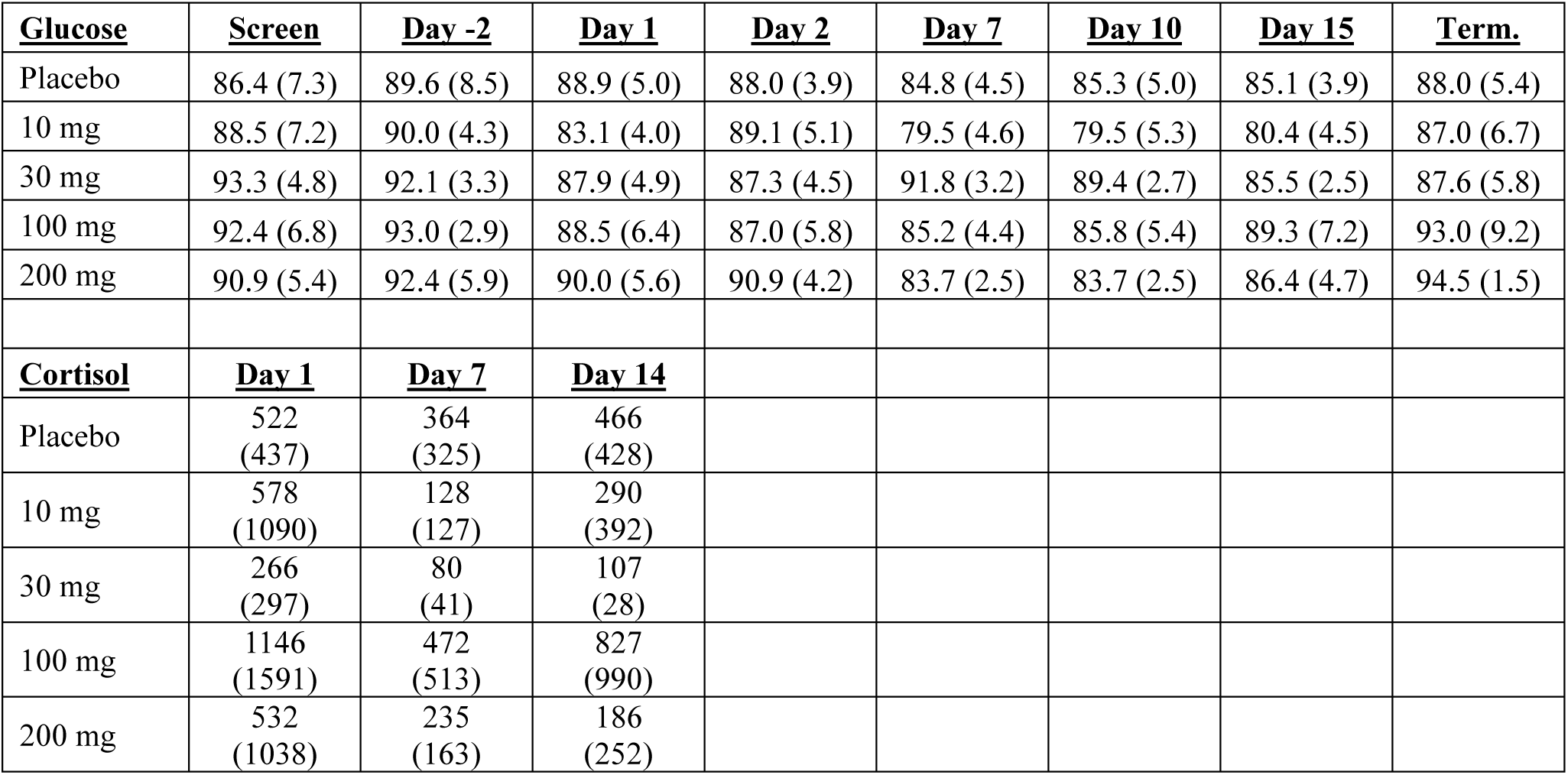
Glucose and Urine Free Cortisol levels for PRX-3140. All laboratory assessments were performed at the clinical site’s certified laboratory utilizing that laboratory’s normal ranges. Blood samples were collected at Screening, Day -2, 1, 2, 7, 10, 15 and following termination shown as mg/dL. Glucose expressed as arithmetic mean (SD). Urine Free Cortisol levels were collected pre-dose at Day 1, 7, and 14 shown as nmol/L. Urine Free Cortisol expressed as arithmetic mean (SD).

Urine free cortisol levels in the 14-day study at 10, 30, 100 and 200 mg dose levels of PRX-3140 compared to placebo demonstrate a reduction at 7 and 14 days compared to initial Day 1 (Figure 6). In particular, 10 mg PRX-3140 demonstrated a reduction from -450 vs. 159 nmol/L at Day 7 and -288 vs. -57 nmol/L at Day 14 for 10mg PRX-3140 vs. placebo, respectively (Table 3). Because of the small patient number (n=8) and interpatient variability, the reduction for placebo vs. 10 mg PRX-3140 at Days 7 and 14 were not statistically significant (p=0.15 and 0.27, respectively). 100 mg PRX-3140 demonstrated a higher reduction from -674 vs. 159 nmol/L at Day 7 and -318 vs. -57 nmol/L at Day 14 for 100 mg PRX-3140 vs. placebo, respectively, but baseline urine free cortisol at Day 1 was higher for 100 mg PRX-3140. 200 mg PRX-3140 demonstrated a reduction from -298 vs. 159 nmol/L at Day 7 and -346 vs. -57 nmol/L at Day 14 for 200 mg PRX-3140 vs. placebo, respectively, but accompanied a higher incidence of side effects. The trend for all mean PRX-3140 doses demonstrated lower cortisol levels at Day 7 and 14 compared to placebo.

**Figure 6:**
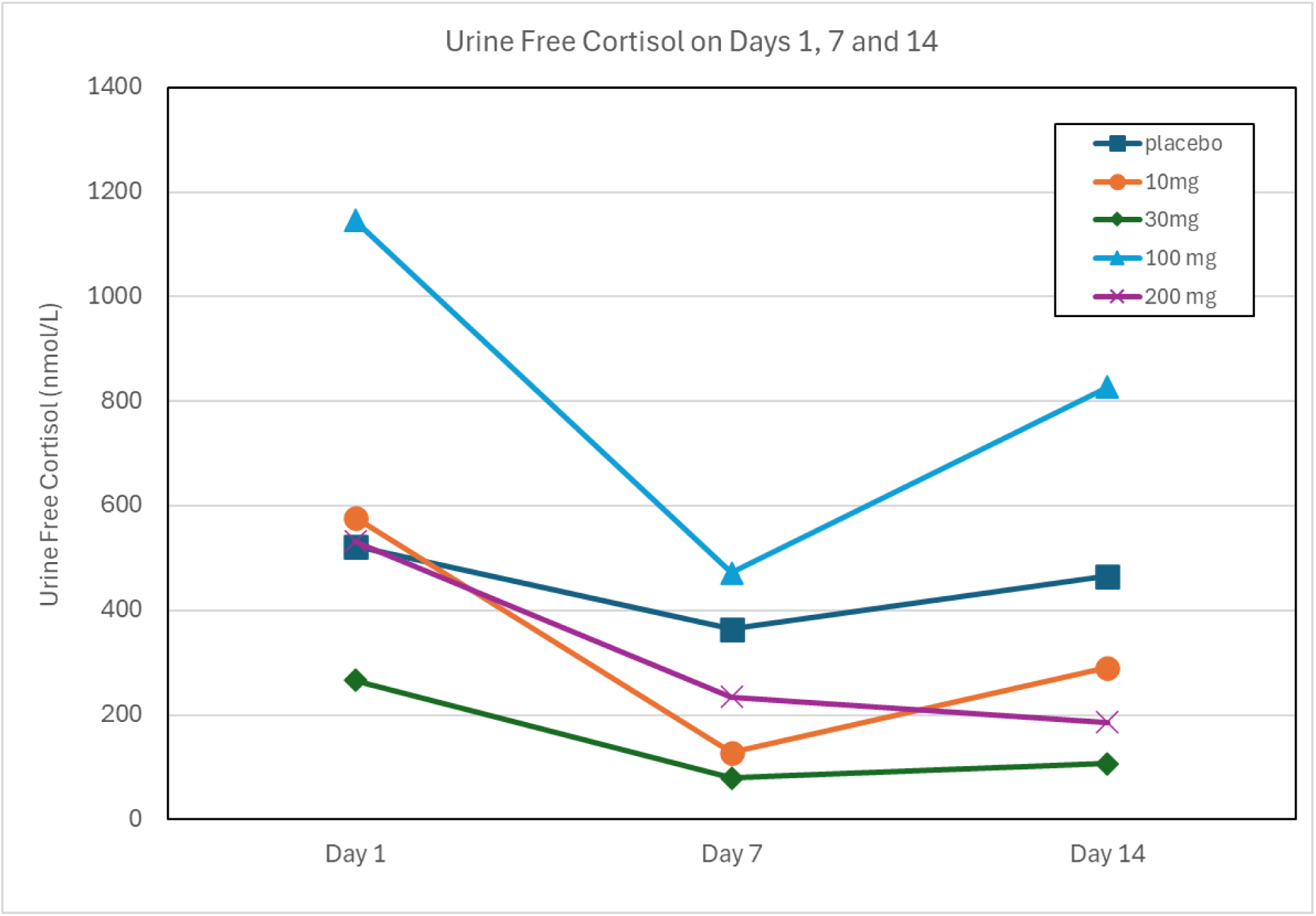
Urine Free Cortisol on Days 7 and 14. Urine Free Cortisol in Subjects pre-dose on Day 1, 7 and Day 14 with oral administration of 10 mg (n=6), 30 mg (n=6), 100 mg (n=6), 200 mg (n=6) PRX-03140 or Placebo (n=8) for 14 days.

## 4 Discussion

Direct acting medications that lower cortisol levels include ketoconazole, metyrapone, mitotane, mifepristone, osilodrostat, and pasireotide, blocking the production or binding to the GR. All of these drugs have notable side effects, including adrenal insufficiency, headache, vomiting, nausea, fatigue, and edema (swelling caused by fluid retention), and with mifepristone abortion. As an agonist of S1R that acts as a chaperone of GR, PRX-3140 has demonstrated GR modulating effects in INS-1 cells and in 14-day clinical studies in healthy adults with low incidence of side effects.

Since the report that S1R knock-out mice demonstrated down-regulation of GR attenuating feedback inhibition of the HPA axis [Di-2017], the potential of S1R agonists to modulate GR-mediated insulin suppression and cortisol levels became of interest. Di et al. presented *in vivo* evidence to show that S1R deficiency in CRF cells attenuates the GR-mediated feedback inhibition of the HPA axis and facilitates the stress response of the HPA axis by down-regulation of PKC signaling to suppress the GR phosphorylation. The results of the present study suggest that S1R activation, with PRX-3140 and NP-18-2 S1R agonists, modulates glucocorticoid insulin suppression and cortisol levels.

In receptor binding assays, PRX-3140 is highly potent (Ki = 22nM) and selective for 5-HT4R with more than 100-fold difference in affinities compared with all other 5-HT receptors tested. PRX-3140 also displays affinity for the S1R (Ki = 79-160nM) and S2R (Ki = 40-100nM) in receptor binding assays. NP-18-2 is a selective agonist for the 5-HT1A with affinity for the S1R but not the S2R. NP-18-3 is a selective antagonist to the 5-HT2B and also demonstrates binding to the S2R (Ki = 42 nM) and S1R (Ki = 100 nM). The three compounds demonstrate selectivity for different 5-HT receptors and differential selectivity for S1R and S2R, with NP-18-2 with the highest S1R selectivity. Binding to over 50 other receptor subtypes was low and did not overlap for any one compound.

Receptor binding assay results cannot reflect activity, and while a cell-based assays monitoring the dissociation of the sigma-1 receptor from its chaperone protein, BiP (also known as GRP78) is used to determine activity [Yano et al., 2018], this assay is prone to variability based on conditions and S1R ligands tested that may complicate interpretation. Therefore, to confirm the S1R activity of PRX-3140, *in vitro* electrophysiology was used as a functional assay. Neuronal excitability on NAc excitability showed a dose dependent reduction in maximum evoked AP firing similar to results with the known S1R antagonist CM304 [Allyene-2023]. Notably, PRX-3140 and NP-18-2 demonstrate S1R antagonist activity at high doses, but reports have also shown antagonists administered at low concentrations can produce the agonist-like effects [Maurice-2021].

Cortisol, an endogenous glucocorticoid hormone produced with a circadian daily rhythm, plays a critical role in the body’s stress response, but prolonged elevation of cortisol levels can lead to various health issues. Cortisol is released by the adrenal cortex in response to adrenocorticotropic hormone (ACTH), which is released by the pituitary in response to corticotropin-releasing hormone (CRH) released by the hypothalamus in response to stress. Chronic stress and high cortisol levels are associated with conditions such as obesity, hypertension, diabetes, and cardiovascular diseases [Abraham-2013]. Dehydroepiandrosterone (DHEA), proposed as the endogenous ligand for the S1R (Ki=15 µM), is the most abundant circulating steroid in the human body produced in the adrenal gland and *de novo* in the central nervous system [Yabuki-2015]. After smoking a cigarette, plasma ACTH and epinephrine increase, followed by cortisol and DHEA reaching peak levels between 30 and 60 minutes after smoking [Mendelson-2005]. The increases in cortisol and DHEA were significantly correlated, and each had a similar half-life after high nicotine cigarette smoking.

Cortisol and DHEA interact with the S1R and can influence the expression and activity of S1R, affecting neuronal health and resilience to stress [Rennekamp-2016]. The S1R is thought to play a protective role in neuronal survival and stress response, and its modulation could offer therapeutic avenues for addressing stress-related disorders [Penke-2018]. Cortisol suppresses insulin and glucagon-like peptide-1 (GLP-1) production in the pancreas and can lead to insulin resistance [Joseph-2022]. The effect of cortisol, DHEA, and the S1R agonist PRX-3140 on insulin production was measured using the INS-1 832/13 rat insulinoma cell line. S1R agonists DHEA, PRX-3140 and NP-18-2 alone moderately increased insulin release at 10 µM, but this increase was not statistically significant. 100 nM cortisol significantly reduced insulin release (48.8 ng/ml of insulin released compared to 117.3 ng/ml exposed to 16.7 mM glucose alone, P-value of <0.0001). If 10 µM PRX-3140 was co-administered with the 100 nM cortisol, the insulin release was significantly increased (74.8 ng/ml, P-value <0.0001). This improvement of insulin release was not seen with DHEA-S or DHEA co-administration. Co-administration of 1 µM DHEA-S with 100 nM cortisol yielded no difference from 100 nM cortisol alone. Co-administration of 10 nM DHEA and 100 nM cortisol drops the amount of insulin released (37.0 ng/ml) when compared to 100 nM cortisol alone (48.8 ng/ml, P value = 0.0015). Similar effects were observed when cells were exposed to dexamethasone, with 10 µM PRX-3140 with the 10 nM Dex producing significantly more insulin than Dex alone. Dex+NP-18-2 demonstrated similar insulin release recovery to PRX-3140, however, Dex+NP-18-3 (S2R ligand) did not, potentially because the concentration tested or 5-HT2 / S2R activity.

5-HT1 and 5-HT4 agonists are known to interact with the HPA. The S1R is proposed as a contributor in modulation of glucocorticoid insulin suppression and cortisol levels. PRX-3140 was shown to lower glucose and cortisol levels at low 10 mg daily doses up to 14 days. In a phase I clinical study NP-18-2 demonstrated lower glucose levels at low oral doses and cortisol lowering effects 1-6 hours after administration (unpublished results). PRX-3140 and NP-18-2, both S1R ligands in the 100 nm range, demonstrated similar ability to improve insulin release and lower cortisol levels at low doses but variable effects at higher doses in clinical studies suggesting a biphasic dose response relationship. Although 5-HT1 and 5-HT4 receptors are known to modulate insulin release and cortisol, the biphasic dose response suggests that S1R activity is a factor. Further clinical studies are warranted to study these effects.

## 5. Data Availability Statement

The raw data supporting the conclusions of this article will be made available by the corresponding author, without undue reservation.

## 7. Author Contributions

BH performed experiments in INS-1 832/13 rat insulinoma cells, completed data analysis, manuscript writing, and production of final figures. JM, AA, JST, SWH, and CJF performed and assisted with the in vitro electrophysiology experiments, development of software tools for data analysis and manuscript editing. JDT wrote the initial version of the manuscript, interpreted clinical data and pharmacokinetics, assisted with direction of the project and manuscript editing.

## 9. Acknowledgements

Authors thank Gunther Hochhaus, Ph.D., Professor, Dept. of Pharmaceutics, College of Pharmacy, Univ. of Florida, for review.

## References

Johnson DE, Drummond E, Grimwood S, Sawant-Basak A, Miller E, Tseng E, McDowell LL, Vanase-Frawley MA, Fisher KE, Rubitski DM, Stutzman-Engwall KJ, Nelson RT, Horner WE, Gorczyca RR, Hajos M, Siok CJ. The 5-hydroxytryptamine4 receptor agonists prucalopride and PRX-03140 increase acetylcholine and histamine levels in the rat prefrontal cortex and the power of stimulated hippocampal θ oscillations. J Pharmacol Exp Ther. 2012 Jun;341(3):681–91. doi: 10.1124/jpet.112.192351. Epub 2012 Mar 9. PMID: 22408061

Nguyen L, Lucke-Wolds BP, Mookerjee S, Kaushal N, Matsumoto RR. Sigma-1 Receptors and Neurodegenerative Diseases: Towards a Hypothesis of Sigma-1 Receptors as Amplifiers of Neurodegeneration and Neuroprotection. Adv Exp Med Biol. 2017;964:133–152. doi: 10.1007/978-3-319-50174-1_10

van Waarde AV, Ramakrishnan NK, Rybczynska AA, Elsinga PH, Ishiwata K, Nijholt IM, Luiten PG, Dierckx RA. The cholinergic system, sigma-1 receptors and cognition. Behavioural Brain Research. Volume 221, Issue 2, 10 August 2011, Pages 543-554

Walter RB, Hoofnagle AN, Lanum SA, Collins SJ. Acute, life-threatening hypoglycemia associated with haloperidol in a hematopoietic stem cell transplant recipient. Bone Marrow Transplant. 2006 Jan;37(1):109–10. doi: 10.1038/sj.bmt.1705187. PMID: 16247427

Marquard J, Otter S, Welters A, Stirban A, Fischer A, Eglinger J, Herebian D, Kletke O, Klemen MS, Stožer A, Wnendt S, Piemonti L, Köhler M, Ferrer J, Thorens B, Schliess F, Rupnik MS, Heise T, Berggren PO, Klöcker N, Meissner T, Mayatepek E, Eberhard D, Kragl M, Lammert E. Characterization of pancreatic NMDA receptors as possible drug targets for diabetes treatment. Nat Med. 2015 Apr;21(4):363–72. doi: 10.1038/nm.3822. Epub 2015 Mar 16. PMID: 25774850

Kavitha J, Arumugam V, Prabakaran E. Influence of Pentazocine on Serum Lipids and Selected Enzymes of Male Wistar Rats J. Clin. Biochem. Nutr., 27, 1–7, 1999.

Paniagua JA. Nutrition, insulin resistance and dysfunctional adipose tissue determine the different components of metabolic syndrome. World J Diabetes. 2016 Nov 15;7(19):483–514. doi: 10.4239/wjd.v7.i19.483. PMID: 27895819

Di T, Zhang S, Hong J, Zhang T, Chen L. Hyperactivity of Hypothalamic-Pituitary-Adrenal Axis Due to Dysfunction of the Hypothalamic Glucocorticoid Receptor in Sigma-1 Receptor Knockout Mice. Front Mol Neurosci. 2017 Sep 6;10:287. doi: 10.3389/fnmol.2017.00287. eCollection 2017. PMID: 28932185

Cheng YC, Prusoff WH. Relationship between the inhibition constant (KI) and the concentration of inhibitor which causes 50 per cent inhibition (I50) of an enzymatic reaction. Biochemical Pharmacology. Volume 22, Issue 23, 1 December 1973, Pages 3099-3108

Scala F et al. Environmental Enrichment and Social Isolation Mediate Neuroplasticity of Medium Spiny Neurons through the GSK3 Pathway. Cell Reports. 2018 Apr 10;23(2):555–567. doi: 10.1016/j.celrep.2018.03.062. PMID: 29642012

Tapia CM, Folorunso O, Singh AK, McDonough K, Laezza F. Effects of Deltamethrin Acute Exposure on Nav1.6 Channels and Medium Spiny Neurons of the Nucleus Accumbens. Toxicology. 2020 Jul:440:152488. doi: 10.1016/j.tox.2020.152488. Epub 2020 May 6. PMID: 32387285

Schindelin J, Arganda-Carreras I, Frise E, Kaynig V, Longair M, Pietzsch T, Preibisch S, Rueden C, Saalfeld S, Schmid B, Tinevez J-Y, White DJ, Hartenstein V, Eliceiri K, Tomancak P, Cardona A. Fiji: an open-source platform for biological-image analysis. 2012. Nat Methods 9:676–682.

Belleau ML, Warren RA. Postnatal Development of Electrophysiological Properties of Nucleus Accumbens Neurons. 2000. J Neurophysiol 84:2204–2216.

Cao J, Dorris DM, Meitzen J. Neonatal Masculinization Blocks Increased Excitatory Synaptic Input in Female Rat Nucleus Accumbens Core. 2016. Endocrinology 157:3181–3196.

Willett JA, Johnson AG, Vogel AR, Patisaul HB, McGraw LA, Meitzen J. Nucleus accumbens core medium spiny neuron electrophysiological properties and partner preference behavior in the adult male prairie vole, Microtus ochrogaster. 2018. J Neurophysiol 119:1576–1588.

Aceto G, Nardella L, Lazzarino G, Tavazzi B, Bertozzi A, Nanni S, Colussi C, D’Ascenzo M, Grassi C. Acute restraint stress impairs histamine type 2 receptor ability to increase the excitability of medium spiny neurons in the nucleus accumbens. 2022. Neurobiol Dis 175:105932.

Harden SW, pyabf 2.3.7. Available at: https://pypi.org/project/pyabf. 2022.

Marino M, Misuri L, Brogioli D. A new open source software for the calculation of the liquid junction potential between two solutions according to the stationary Nernst-Planck equation. Arxiv. arXiv:1403.3640 [physics.chem-ph] 2014.

Golowasch J, Thomas G, Taylor AL, Patel A, Pineda A, Khalil C, Nadim F. Membrane Capacitance Measurements Revisited: Dependence of Capacitance Value on Measurement Method in Nonisopotential Neurons. 2009. J Neurophysiol 102:2161–2175.

Cirino TJ, Harden SW, McLaughlin JP, Frazier CJ. Region-specific effects of HIV-1 Tat on intrinsic electrophysiological properties of pyramidal neurons in mouse prefrontal cortex and hippocampus. 2020. J Neurophysiol 123:1332–1341.

Shen W, Hernandez-Lopez S, Tkatch T, Held JE, Surmeier DJ. Kv1.2-Containing K+ Channels Regulate Subthreshold Excitability of Striatal Medium Spiny Neurons. 2004. J Neurophysiol 91:1337–1349.

Hohmeier HE, Mulder H, Chen G, Henkel-Rieger R, Prentki M, Newgard CB. Isolation of INS-1-derived cell lines with robust ATP-sensitive K+ channel-dependent and -independent glucose-stimulated insulin secretion. Diabetes. 2000 Mar;49(3):424–30. doi: 10.2337/diabetes.49.3.424. PMID: 10868964

Alleyne AR. A Mechanistic Interrogation of the Sigma-1 Receptor as a Potential Target for Psychostimulant Use Disorder. Dissertation. 2023. https://original-ufdc.uflib.ufl.edu/UFE0059777/00001

Abraham SB, Rubino D, Sinaii N, Ramsey S, Nieman LK. Cortisol, obesity and the metabolic syndrome: A cross-sectional study of obese subjects and review of the literature. Obesity (Silver Spring). 2013 Jan;21(1):E105–E117. doi: 10.1002/oby.20083. PMID: 23505190

Suksri K, Semprasert N, Limjindaporn T, Yenchitsomanus PT, Kooptiwoot S, Kooptiwut S. Cytoprotective effect of genistein against dexamethasone-induced pancreatic beta-cell apoptosis. Sci Rep. 2022 Jul 28;12(1):12950. doi: 10.1038/s41598-022-17372-z. PMID: 35902739

Maurice T. Bi-phasic dose response in the preclinical and clinical developments of sigma-1 receptor ligands for the treatment of neurodegenerative disorders. Expert Opinion on Drug Discovery, 16:4, 373–389, DOI: 10.1080/17460441.2021.1838483

Yabuki Y, Shinoda Y, Izumi H, Ikuno T, Shioda N, Fukunaga K. Dehydroepiandrosterone administration improves memory deficits following transient brain ischemia through sigma-1 receptor stimulation. Brain Res. 2015 Oct 5;1622:102–13. doi: 10.1016/j.brainres.2015.05.006. Epub 2015 Jun 25. PMID: 26119915

Rennekamp AJ, Huang XP, Wang Y, Patel S, Lorello PJ, Cade L, Gonzales AP, Yeh JR, Caldarone BJ, Roth BL, Kokel D, Peterson RT. Sigma-1 receptor ligands control a switch between passive and active threat responses. Nat Chem Biol. 2016 May 30;12(7):552–558. doi: 10.1038/nchembio.2089. PMID: 27239788.

Penke B, Fülöp L, Szűcs M, Frecska E. The Role of Sigma-1 Receptor, an Intracellular Chaperone in Neurodegenerative Diseases. Curr Neuropharmacol. 2018 Jan;16(1):97–116. PMID: 28554311

Joseph AM and Janssen JL. New Insights into the Role of Insulin and Hypothalamic-Pituitary-Adrenal (HPA) Axis in the Metabolic Syndrome. Int J Mol Sci. 2022 Jul 25;23(15):8178. doi: 10.3390/ijms23158178. PMID: 35897752

Yano H, Bonifazi A, Xu M, Guthrie DA, Schneck SN, Abramyan AM, Fant AD, Hong WC, Newman AH, Shi L. Pharmacological Profiling of Sigma 1 Receptor Ligands by Novel Receptor Homomer Assays. Neuropharmacology. 2018 Jan 31;133:264–275. doi: 10.1016/j.neuropharm.2018.01.042. PMID: 29407216.

